# Detection of SARS-CoV-2 RNA Throughout Wastewater Treatment Plants and A Modeling Approach to Understand COVID-19 Infection Dynamics in Winnipeg, Canada

**DOI:** 10.1101/2021.10.26.21265146

**Authors:** Kadir Yanaç, Adeola Adegoke, Liqun Wang, Qiuyan Yuan, Miguel Uyaguari

## Abstract

Although numerous studies have detected SARS-CoV-2 in wastewater and attempted to find correlations between the concentration of SARS-CoV-2 and the number of cases, no consensus has been reached on sample collection and processing, and data analysis. Moreover, the fate of SARS-CoV-2 in wastewater treatment plants is another issue, specifically regarding the discharge of the virus into environmental settings and the water cycle. The current study monitored SARS-CoV-2 in influent and effluent wastewater samples with three different concentration methods and sludge samples over six months (July to December 2020) to compare different virus concentration methods, assess the fate of SARS-CoV-2 in wastewater treatment plants, and describe the potential relationship between SARS-CoV-2 concentrations in influent and infection dynamics. Skimmed milk flocculation (SMF) resulted in higher recoveries (15.27% ± 3.32%) of an internal positive control, Armored RNA, and higher positivity rate of SARS-CoV-2 in samples compared to ultrafiltration methods employing a prefiltration step to eliminate solids. Our results suggested that SARS-CoV-2 may predominate in solids and therefore, concentration methods focusing on both supernatant and solid fractions may result in better recovery. SARS-CoV-2 was detected in influent and primary sludge samples but not in secondary and final effluent samples, indicating a significant reduction during primary and secondary treatments. SARS-CoV-2 was first detected in influent on September 30^th^, 2020. A decay-rate formula was applied to estimate initial concentrations of late-processed samples with SMF. A model based on shedding rate and new cases was applied to estimate SARS-CoV-2 concentrations and the number of active shedders. Inferred sensitivity of observed and modeled concentrations to the fluctuations in new cases and test-positivity rates indicated a potential contribution of newly infected individuals to SARS-CoV-2 loads in wastewater.

## 1. Introduction

After the initial emergence of COVID-19 in December 2019 in Wuhan, China, the disease has rapidly spread and caused significant health, economic and social burden worldwide. Considering the rapid transmission and spread of COVID-19 and the subsequent emergence of new waves and variants, surveillance of the disease is vital to early predict and control outbreaks in communities. Standard diagnostic testing among populations is challenging due to the time and cost to test massive numbers of individuals. Moreover, undiagnosed and unreported asymptomatic and mildly symptomatic cases of COVID-19 constitute a significant proportion of infections (Bi et al., 2021; Day, 2020; Ing et al., 2020), and this increases the complexity of assessing the true scale of a community outbreak. With the first detection of SARS-CoV-2 in urban wastewater reported in the Netherlands in March 2020, when number of cases were very low (and even zero in some locations (Medema et al., 2020a)), wastewater-based epidemiology (WBE) has emerged as an additional surveillance tool to mitigate these challenges and provide rapid information to government agencies, civil society organizations, and private or public utilities to determine the effectiveness of public health control measures and the required level of societal restrictions (Bivins et al., 2020b; Hill et al., 2020; Thompson et al., 2020). Moreover, the existence of SARS-CoV-2 in wastewater treatment plants (WWTPs) is concerning because of its health risks to the workers and its potential dissemination into the environment through discharge.

WBE has been used as a surveillance and predictive tool to provide real-time information on the usage of illegal drugs (Bishop et al., 2020; Sulej-Suchomska et al., 2020), and the prevalence of viral (Bisseux et al., 2018; Ivanova et al., 2019; McCall et al., 2020; Nakamura et al., 2015) and bacterial (Diemert and Yan, 2020; Yan et al., 2018) diseases. Presence of SARS-CoV-2 gene fragments in the feces of both symptomatic and asymptomatic infected individuals (Gupta et al., 2020; Jones et al., 2020; Pan et al., 2020) makes wastewater surveillance of COVID-19 a unique epidemiological tool to monitor infection trends in communities and support public health interventions. Indeed, since the onset of COVID-19, many researchers have reported the detection and quantitation of SARS-CoV-2 in wastewater (Ahmed et al., 2020a; D’Aoust et al., 2021; Graham et al., 2021; Medema et al., 2020a; Westhaus et al., 2021). Several potential uses of the generated data include tracking trends temporally to project future infection trajectories (Medema et al., 2020b) and estimating COVID-19 prevalence in a community based on SARS-CoV-2 concentration in wastewater (Ahmed et al., 2020a; Gerrity et al., 2021; Wu et al., 2022). Moreover, surveillance of primary sewage sludge and wastewater solids, specifically the partition of enveloped viruses to the wastewater solids (Ye et al., 2016), has also been reported as an alternative to wastewater influent surveillance (Graham et al., 2021; Peccia et al., 2020). In fact, processing sludge and solids samples using reverse-transcriptase quantitative polymerase chain reaction (RT-qPCR) (Peccia et al., 2020) is not as labor-intensive as processing wastewater influent samples, in which viral particles are concentrated either using filtration methods or organic flocculation methods (Ahmed et al., 2020c; Barril et al., 2021). However, regardless of the surveillance type, there are still some methodological questions on the cumulative interpretation of data generated from wastewater analysis and epidemiological data together.

Our knowledge of the fate of coronaviruses in environmental compartments is very limited (Carducci et al., 2020). Westhaus et al. (2021) detected SARS-CoV-2 in treated wastewater, indicating its potential distribution into aquatic ecosystems, while Hasan et al. (2021) could not detect SARS-CoV-2 in treated wastewater. Detection of SARS-CoV-2 in wastewater solids and sludge (Balboa et al., 2020; D’Aoust et al., 2021; Graham et al., 2021; Peccia et al., 2020) might indicate the predominance of SARS-CoV-2 in solids and sludge line as the primary removal mechanism in WWTPs. Thus, concentration methods focusing on the recovery from both solids and supernatant may better approach the true concentrations of SARS-CoV-2 in wastewater (Chik et al., 2021). Different wastewater treatment processes, sewage characteristics, and climate conditions may affect the fate of SARS-CoV-2 in WWTPs. More in-depth investigations will provide better insights into the fate of SARS-CoV-2 throughout WWTPs and its circulation in the water cycle.

This study monitored SARS-CoV-2 concentrations using raw composite wastewater samples from three WWTPs in Winnipeg over six months (July to December 2020), characterized by increasing COVID-19 incidence and prevalence. We applied three different concentration methods to process raw wastewater samples. SARS-CoV-2 concentrations in raw wastewater were analyzed for the population-wide infection dynamics in Winnipeg. We used a back trajectory modeling approach to estimate the initial concentration of SARS-CoV-2 genes in the late processed wastewater samples. A second model was also applied to estimate the concentration of SARS-CoV-2 based on reported cases and shedding rate. Furthermore, we also monitored SARS-CoV-2 concentrations in the effluent and primary sludge samples to understand the fate of SARS-CoV-2 in WWTPs.

## 2. Materials and Methods

### 2.1. Sample Collection

One-liter of 24-h composite raw wastewater samples were collected from three major WWTPs in Winnipeg, Canada, between July 8 and December 15, 2020. These WWTPs are the North End Sewage Treatment Plant (NESTP), South End Sewage Treatment Plant (SESTP), and West End Sewage Treatment Plant (WESTP). Primary sludge (50 mL), secondary effluent (1 L), and final effluent (1 L) samples were also collected during this period. Detailed information on sampling dates, sample types, and treatment processes of WWTPs and their design capacities is provided in Table S1 and Table S2, respectively. The operators provided daily flowrate (Fig. S1) and wastewater characteristics data. Wastewater and sludge samples were transported to the laboratory in amber HDPE bottles in an icebox after sampling and processed within 24 hours.

### 2.2. Virus Concentration Assays

Three different virus concentration methods were applied: an ultrafiltration method at two different centrifugation speeds and an organic flocculation method.

#### Ultrafiltration

Samples were processed within 24 hours of collection. Raw wastewater, and secondary and final effluent samples (120 mL each) were filtered through cheesecloth and low-protein binding 0.45 and 0.2 µm 47-mm Supor-200 membrane disc filters (Pall Corporation, Ann Harbor, MI), respectively, to remove large particles, sediments, eukaryotes, and bacteria (Uyaguari-Diaz et al., 2016). Solids retained on the filters were stored at –20 °C.

##### Sequential ultrafiltration at 3000g (UF-3K x g)

A total of 120 mL of the sample was first concentrated, at 3000 g for 30 minutes by loading 60 mL of the sample twice, to approximately 5 mL using Jumbosep Centrifugal Device, 30-kDa (Pall Corporation, Ann Harbor, MI, USA). Then, the 5 mL concentrate was further concentrated down at 3000 g for 30 minutes using Microsep Advance Centrifugal Device, 30-kDa, (Pall Corporation, Ann Harbor, MI, USA). The final volume of the concentrate varied between 500 and 1200 µL.

##### Sequential ultrafiltration at 7500g (UF-7.5K x g)

To get smaller final volumes and as per the manufacturer’s recommendation, centrifugation speed for Microsep was changed from 3000 g to 7500 g from October 28^th^, 2020.

Samples collected between October 28^th^ and December 15^th^ were processed with both UF-3K x g and UF-7.5K x g methods.

#### Organic Flocculation

The samples collected between November 16^th^ and December 15^th^ were additionally concentrated by applying skimmed milk flocculation (SMF) protocol (Calgua et al., 2008) even though a long time (Table 1) had passed since the sample collection. Briefly, 0.5 g of skim milk powder (Difco Laboratories, Sparks, MD, USA) was dissolved in 50 mL synthetic seawater (Sigma-Aldrich, St. Louis, MO, USA) to obtain 1% (w/v) skimmed milk solution and pH of the solution was carefully adjusted with 1M HCl to 3.5. Five mL of skimmed milk solution was added to 500 mL raw wastewater samples. pH of samples was also previously adjusted to 3.5 with 1M HCl to obtain a final concentration of skimmed milk at 0.01% (w/v). Samples were stirred for eight hours, and flocs were allowed to settle for another eight hours at room temperature. Supernatants were carefully removed using serological pipets without disturbing the settled flocs. A final volume of 50 mL containing the flocs was transferred to centrifuge tubes and centrifuged at 8000 x g for 30 minutes. Pellets were carefully scraped using a sterilized spatula, and the remaining pellets in the tubes were resuspended in 250 µL of 0.2M sodium phosphate buffer with a pH 7.5 (Alfa Aesar, Ottawa, ON, Canada) and transferred to 1.5 mL Eppendorf tubes.

**Table 1.** Delay in sample processing collected from NESTP, SESTP, and WESTP

| Sampling date | Delay in sample processing (days) |
| --- | --- |
| November 16, 2020 | 75 |
| November 18, 2020 | 73 |
| December 1, 2020 | 60 |
| December 8, 2020 | 53 |
| December 15, 2020 | 46 |

##### Recovery Efficiency

Total recovery for ultrafiltration and SMF method workflows were determined by spiking 5×10^4^ copies of Armored RNA Quant IPC-1 Processing Control (Asuragen, Austin, TX, USA) (Alygizakis et al., 2021; Eveleigh et al., 2019; Hietala and Crossley, 2006) into six raw wastewater samples from North End Sewage Treatment Plant (NESTP), South End Sewage Treatment Plant (SESTP) and West End Sewage Treatment Plant (WESTP) in Winnipeg for each virus concentration method and then stirring and inoculating the samples at 4°C for 30 minutes. Recovery efficiency percentage was calculated by dividing the recovered concentration by the spiked concentration.

### 2.3. Viral RNA Extraction

Viral concentrates from wastewater samples and recovery efficiency assays were processed using the RNeasy PowerMicrobiome Kit (Qiagen Sciences, Inc., Germantown, MD, USA). Phenol:Chloroform: Isoamyl Alcohol 25:24:1 (Invitrogen, Carlsbad, CA, USA) and β-mercaptoethanol (Fisher Scientific, Fair Lawn, NJ, USA) were also added during extraction according to the manufacturer’s instructions to improve extraction efficiency. 300 µL of sludge samples were added directly to the beading tubes to extract viral RNA from sludge using the same extraction kit and following the same instructions. Finally, RNA was eluted in 50 µL of elution buffer. On the other hand, viral RNA was extracted from wastewater solids using MagMAX Microbiome kit (Thermo Fisher, Waltham, MA, USA) following the manufacturer’s instructions that included the addition of Proteinase K. The RT-qPCR analysis was conducted following the extraction, and extracts were stored at −80 °C.

### 2.4. RT-qPCR Analysis

In this study, N1 and N2 primers/probe sets (Integrated DNA Technologies, Inc., Coralville, IA, USA), each targeting a different region of the Nucleocapsid (N) gene of SARS-CoV-2 (CDC, 2020), were used. Each 10-μl RT-qPCR mixture consisted of 10 μl of 2.5 µL of 4X TaqMan Fast Virus 1-Step Master Mix (Life Technologies, Carlsbad, CA, USA), 400 nM each primer, 250 nM probe, 2.5 μl of the template and ultrapure DNAse/RNAse free distilled water (Promega Corporation, Fitchburg, WI, USA). Calibration curves for quantifying N1 and N2 specific assays were obtained using six 10-fold dilutions (ranging from 2.0 to 2.0E+05) of the 2019-nCoV_N_Positive Control plasmid DNA (Integrated DNA Technologies, Inc., Coralville, IA, USA). Armored RNA was quantified using 10-fold dilutions of synthetic single-stranded DNA (Integrated DNA Technologies, Inc., Coralville, IA, USA). Primers and probe sets (Integrated DNA Technologies, Inc., Coralville, IA, USA) used to detect and quantify the Armored RNA are given in Table S3. Calibration curves were obtained for each RT-qPCR run. Negative controls were also included in each qPCR run. Thermal cycling reactions were performed at 50 °C for 5 minutes, followed by 45 cycles of 95 °C for 10 seconds and 60 °C for 30 seconds on a QuantStudio 5 Real-Time PCR System (Life Technologies, Carlsbad, CA, USA). Following recommendations by the CDC’s protocol for detection of SARS-CoV-2 (2020) if the C_T_ was <40, samples were considered SARS-CoV-2 positive.

The concentration of SARS-CoV-2 in wastewater in Winnipeg was normalized by dividing daily total SARS-CoV-2 load in wastewater by the total daily wastewater flow rate (eqn. 1). The normalization was implemented for the three WWTPs using eqn. 1, where NC is normalized concentration, C represents the SARS-CoV-2 concentration, and WF indicates the daily wastewater flow.

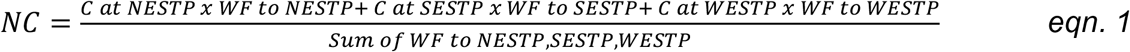

### 2.5. Model 1: Back Trajectory Modeling

We used SMF to concentrate SARS-CoV-2 from the raw wastewater samples stored at 4 °C. The delay times in sample processing with SMF approach are provided in Table 1.

To estimate the initial concentration of SARS-CoV-2 genes on the sampling day, a model was derived based on the previously reported decay rate constants for N1 (Ahmed et al., 2020b) and N2 (Hokajärvi et al., 2021a) genes at 4 °C in untreated wastewater (Table 2). Both Ahmed et al. (2020b) and Hokajärvi et al. (2021a) linearized the observed RNA concentrations using the natural logarithm (ln)-transformation of the normalized concentrations (eqn. 2). The decay rate (k) for N2 was re-calculated as 0.063 (R^2^=0.99) in units per day by linear regression using the data reported by Hokajärvi et al. (2021a), while the decay rate (k) reported by Ahmed et al. (2020b) was used for N1 assay.

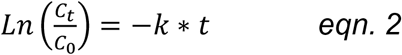

**Table 2.** Reported decay rate characteristics of the SARS-CoV-2 in untreated wastewater at 4°C

| Assay | Decay rate (k) | R<sup>2</sup> | T<sub>50</sub> (days) | T<sub>90</sub> (days) | T<sub>99</sub> (days) | Reference |
| --- | --- | --- | --- | --- | --- | --- |
| N1 | 0.084 ± 0.013 | 0.79 |  | 27.8 ± 4.45 |  | (Ahmed et al., 2020b) |
| N2 | 0.06 ± 0.0 | 0.99 | 11 | 36 | 73 | (Hokajärvi et al., 2021a) |

Where C_t_ is the concentration at time t, C_o_ represents the initial concentration, and k indicates the decay rate. A log-linear model was derived from eqn. 2 for the back-calculation of N1 and N2 concentrations (gene copies) as shown in eqn. 3, where C_t_ is the concentration of N1 or N2 genes at time t, C_0_ is the initial concentration of N1 or N2 genes at the sampling day, k is the decay rate of N1 or N2 genes, and t is the delay in processing the samples.

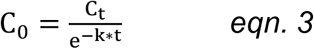

### 2.6. Model 2: Estimation of Number of Shedding Cases, and SARS-CoV-2 Concentration

The second model was adopted from Gerrity et al. (2021) with slight changes to estimate SARS-CoV-2 concentration and number of shedding cases as a function of new cases in the city, fecal shedding rate, and daily total wastewater flowrate. The model was rewritten in RStudio since the original script was written in MATLAB. This model assumes the feces production rate and initial fecal shedding rate are 126 g/person-d and 10^8.9^ GC/g feces with a decay rate 10^0.35^ GC/g feces-day, respectively (Gerrity et al., 2021; Wölfel et al., 2020). The ascertainment ratio was assumed as 2 (Gerrity et al., 2021), which means the model will multiply the number of cases by 2. Instead of assuming a constant daily wastewater flow rate, we used daily wastewater flow rates provided by the city of Winnipeg. Based on the shedding rate and shedding decay rate, an infected person is expected to shed SARS-CoV-2 for 26 days with a burst on the initial days of infection.

### 2.7. Epidemiological Data

Information regarding new, cumulative and active cases, and test-positivity rates of COVID-19 was provided by the Manitoba Regional Health Authority (Manitoba, 2021).

## 3. Results and Discussion

### 3.1. Evaluation of SARS-CoV-2 concentration methods

Viral concentration with UF-3K x g resulted in 13.38 ± 9.11 % recovery (Fig. 1) and a final volume varying between 500 and 1200 μL. The high standard deviation could be attributed to the losses between the two-step ultrafiltration process. This two-step process resulted in high final volumes, which require multiple bead tubes and additional use of extraction kit solutions for RNA extraction since bead tubes have a maximum capacity of 300 μL of sample. During extraction, all tubes of the same sample were eluted into the same spinning column to collect total RNA. The use of multiple bead tubes can be another reason for the high variability in the recovery efficiency.

**Fig. 1.**
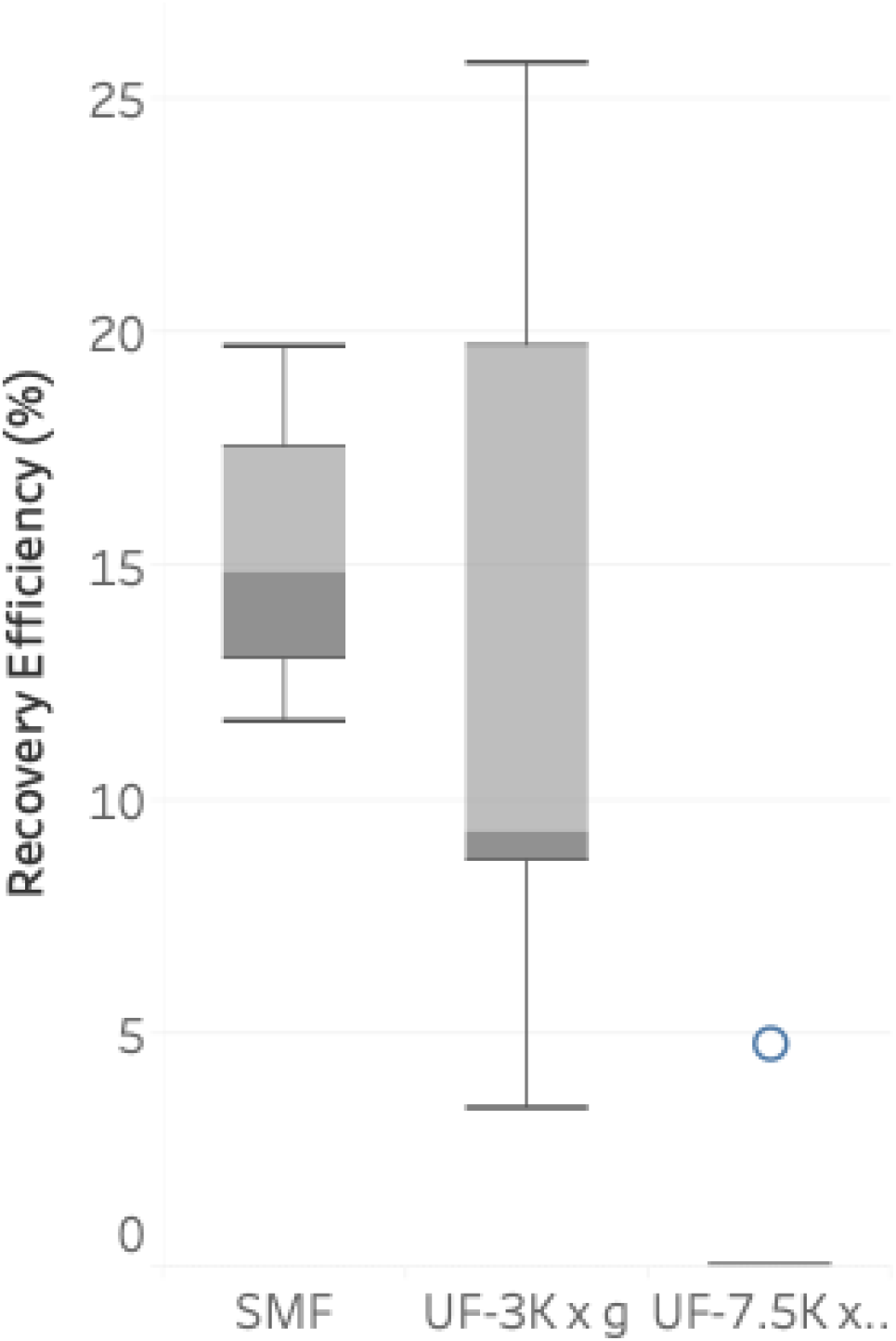
Percent recovery and statistical analysis for each method.

To decrease variability in recovery efficiency potentially caused by the use of multiple bead tubes, the centrifugation speed of Microsep Advance Centrifugal Device was increased from 3000g to 7500g, which generally resulted in final volume less than 400 μL. The first trial with UF-7.5K x g resulted in 4.79% recovery of Armored RNA (Fig. 1). After getting negative or weak SARS-CoV-2 genetic signals in wastewater samples with UF-7.5K x g, further recovery experiments were conducted to determine recovery efficiency at 7500 x g, and but Armored RNA was not recovered. Apparently, increasing the centrifugation speed impacted the recovery, probably caused Armored RNA to pass through or bind onto the filters. Filtration with cheesecloth and low-protein binding 0.45 and 0.2 µm 47-mm membrane filters before ultrafiltration might also be another reason for no detection and weak signals of SARS-CoV-2 due to the elimination of solid particles, which could carry a significant amount SARS-CoV-2 (D’Aoust et al., 2021; Graham et al., 2021; Westhaus et al., 2021).

SMF was later employed to concentrate and detect SARS-CoV-2 in wastewater samples. Using Armored RNA as a control, 15.27% ± 3.32% of recovery was achieved from the spiked wastewater samples (Fig. 1.). SMF has the highest recovery efficiency with the smallest standard deviation whereas UF-7.5K x g has the lowest recovery efficiency with 5 negative samples out of 6 samples.This method did not employ a prefiltration step with cheesecloth and low-protein binding 0.45 and 0.2 µm 47-mm membrane filters. Therefore, the losses due to the elimination of solid particles were minimized and might have been reflected through the higher percentage and lower variation in recovery. Percent recovery values for SMF were comparable to the recovery values reported by Philo et al. (2021). They also reported 30% positivity rate for SARS-CoV-2 with SMF.

### 3.2. *Detection of* SARS-CoV-2 in influent and effluent wastewater and primary sludge

#### 3.2.1. Virus concentration method determines the detection of SARS-CoV-2 in influent samples

Table 3 shows concentrations of SARS-CoV-2 in wastewater and sludge samples collected during the sampling period. Three different concentration methods, namely UF-3K x g, UF-7.5K x g and SMF, were applied (Fig. 2). UF-3K x g and UF-7.5K x g methods required a prefiltration step with 0.45 and 0.2 μm filters to eliminate the effects of solids on ultrafiltration performance and separate viral fraction from bacterial fraction. On the other hand, SMF did not require any prefiltration step, and therefore SARS-CoV-2 particles on solids were also concentrated with this method. All influent and effluent samples were processed using UF-3K x g method (Fig. 2). Influent samples collected between October 28^th^ and December 15^th^ were processed with both UF-3K x g AND UF-7.5K x g methods. They were first concentrated with UF-7.5K x g method, and the negative samples were further processed with UF-3K x g method within 4 days of sampling. As samples were negative when processed with UF-3K x g and UF-7.5K x g methods between November 16^th^ and December 15^th^ (Fig. 2), SMF method was applied delayed (Table 1) to concentrate SARS-CoV-2 from these samples stored at 4 °C. SARS-CoV-2 was detected in all influent samples collected between September 30^th^ and December 15^th^, except October on 28^th^. SARS-CoV-2 was detected only in influent samples collected from SESTP on October 28^th^.

**Table 3.** Concentration of SARS-CoV-2 RNA in influents of three WWTPs. Influent samples collected between November 16^th^ and December 15^th^ (shown in a box) were late processed samples with SMF

| Sampling<br>Date | NESTP |  |  |  |  |  | SESTP |  |  |  |  |  | WESTP |  |  |  |  |  |
| --- | --- | --- | --- | --- | --- | --- | --- | --- | --- | --- | --- | --- | --- | --- | --- | --- | --- | --- |
|  | Influent (GC/L) |  | Primary Sludge |  | Effluent |  | Influent (GC/L) |  | Primary Sludge |  | Effluent |  | Influent (GC/L) |  | Primary Sludge |  | Effluent |  |
|  |  |  | (GC/mL) |  | (GC/L) |  |  |  | (GC/mL) |  | (GC/L) |  |  |  | (GC/mL) |  | (GC/L) |  |
|  | N1 | N2 | N1 | N2 | N1 | N2 | N1 | N2 | N1 | N2 | N1 | N2 | N1 | N2 | N1 | N2 | N1 | N2 |
| 8-Jul | 0 | 0 | - | - | 0 | 0 | 0 | 0 | - | - | 0 | 0 | 0 | 0 | - | - | 0 | 0 |
| 22-Jul | 0 | 0 | - | - | 0 | 0 | 0 | 0 | - | - | 0 | 0 | 0 | 0 | - | - | 0 | 0 |
| 5-Aug | 0 | 0 | - | - | 0 | 0 | 0 | 0 | - | - | 0 | 0 | 0 | 0 | - | - | 0 | 0 |
| 19-Aug | 0 | 0 | - | - | 0 | 0 | 0 | 0 | - | - | 0 | 0 | 0 | 0 | - | - | 0 | 0 |
| 2-Sep | 0 | 0 | - | - | 0 | 0 | 0 | 0 | - | - | 0 | 0 | 0 | 0 | - | - | 0 | 0 |
| 16-Sep | 0 | 0 | - | - | 0 | 0 | 0 | 0 | - | - | 0 | 0 | 0 | 0 | - | - | 0 | 0 |
| 30-Sep | 6,087 | 25,713 | - | - | 0 | 0 | 8,778 | 20,009 | - | - | 0 | 0 | 25,087 | 47,557 | - | - | 0 | 0 |
| 14-Oct | 13,222 | 34,751 | - | - | 0 | 0 | 26,670 | 47,714 | - | - | 0 | 0 | 28,894 | 47,226 | - | - | 0 | 0 |
| 28-Oct | 0 | 0 | - | - | 0 | 0 | 7,544 | 21,084 | - | - | 0 | 0 | 0 | 0 | - | - | 0 | 0 |
| 12-Nov | 3.3E+6 | 2.5E+6 | 7,980 | 4,960 | - | - | 1.2E+6 | 3.4E+6 | 3,140 | 2,740 | - | - | 9.6E+5 | 1.1E+6 | 2,080 | 660 | - | - |
| 16-Nov | 51,400 | 26,668 | 96,200 | 8,860 | - | - | 12,964 | 17,410 | 2,400 | 1,360 | - | - | 16,506 | 4,372 | 0 | 0 | - | - |
| 18-Nov | 16,249 | 55,512 | 8,540 | 6,480 | - | - | 5,843 | 17,412 | 3,280 | 3,040 | - | - | 13,762 | 11,844 | 0 | 380 | - | - |
| 1-Dec | 20,980 | 16,833 | 7,480 | 6,740 | - | - | 11,261 | 6,780 | 3,100 | 2,260 | - | - | 5,519 | 2,214 | 0 | 260 | - | - |
| 8-Dec | 15,722 | 11,463 | 12,580 | 5,860 | - | - | 4,984 | 9,685 | 7,560 | 2,760 | - | - | 9,007 | 5,300 | 820 | 180 | - | - |
| 15-Dec | 20,371 | 13,120 | 3,220 | 1,400 | - | - | 17,209 | 8,188 | 3,020 | 740 | - | - | 5,535 | 5,211 | 5,660 | 2,600 | - | - |

**Table 3.**
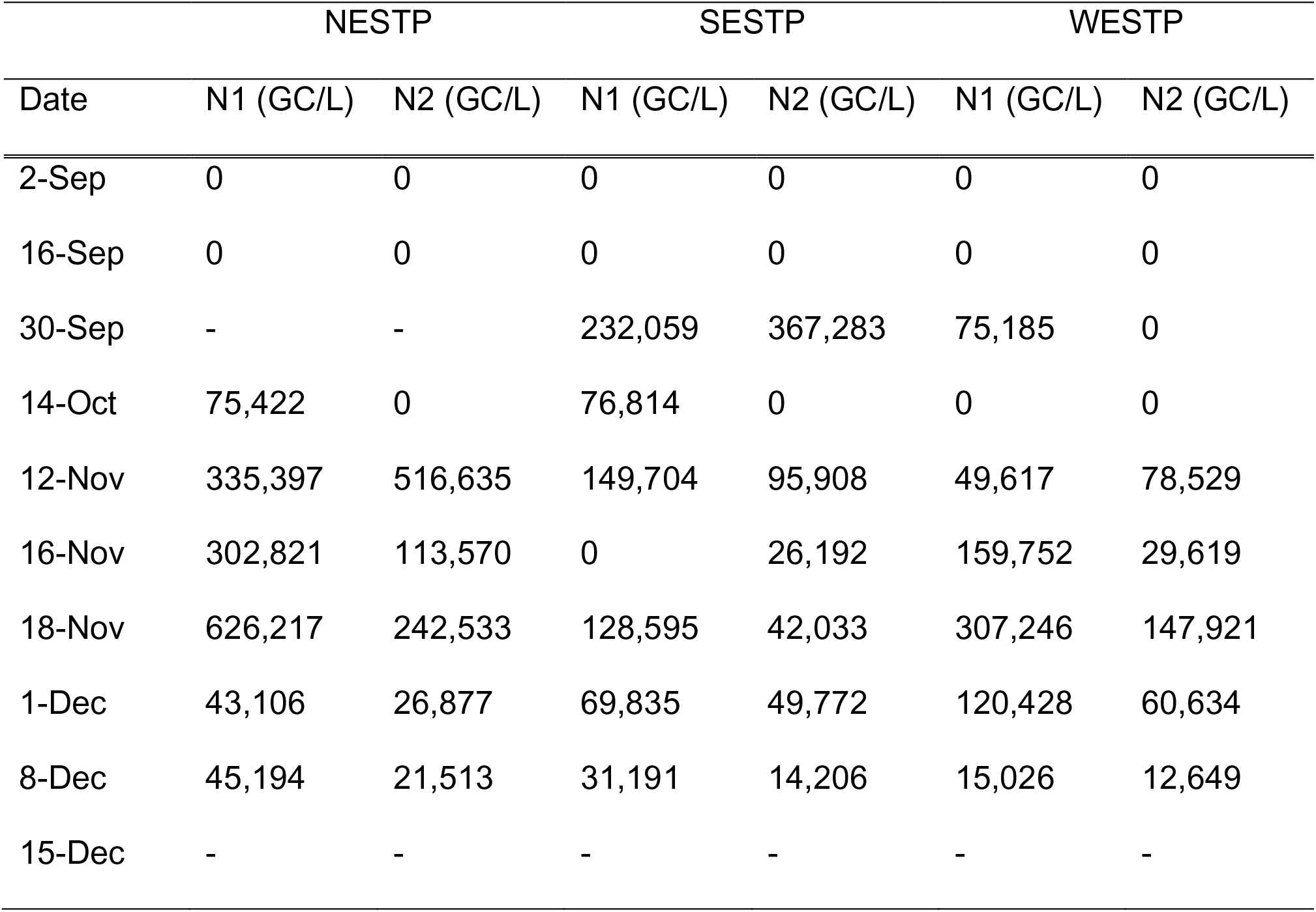
SARS-CoV-2 RNA concentration in influent solids from NESTP, SESTP and WESTP.

**Fig. 2.**
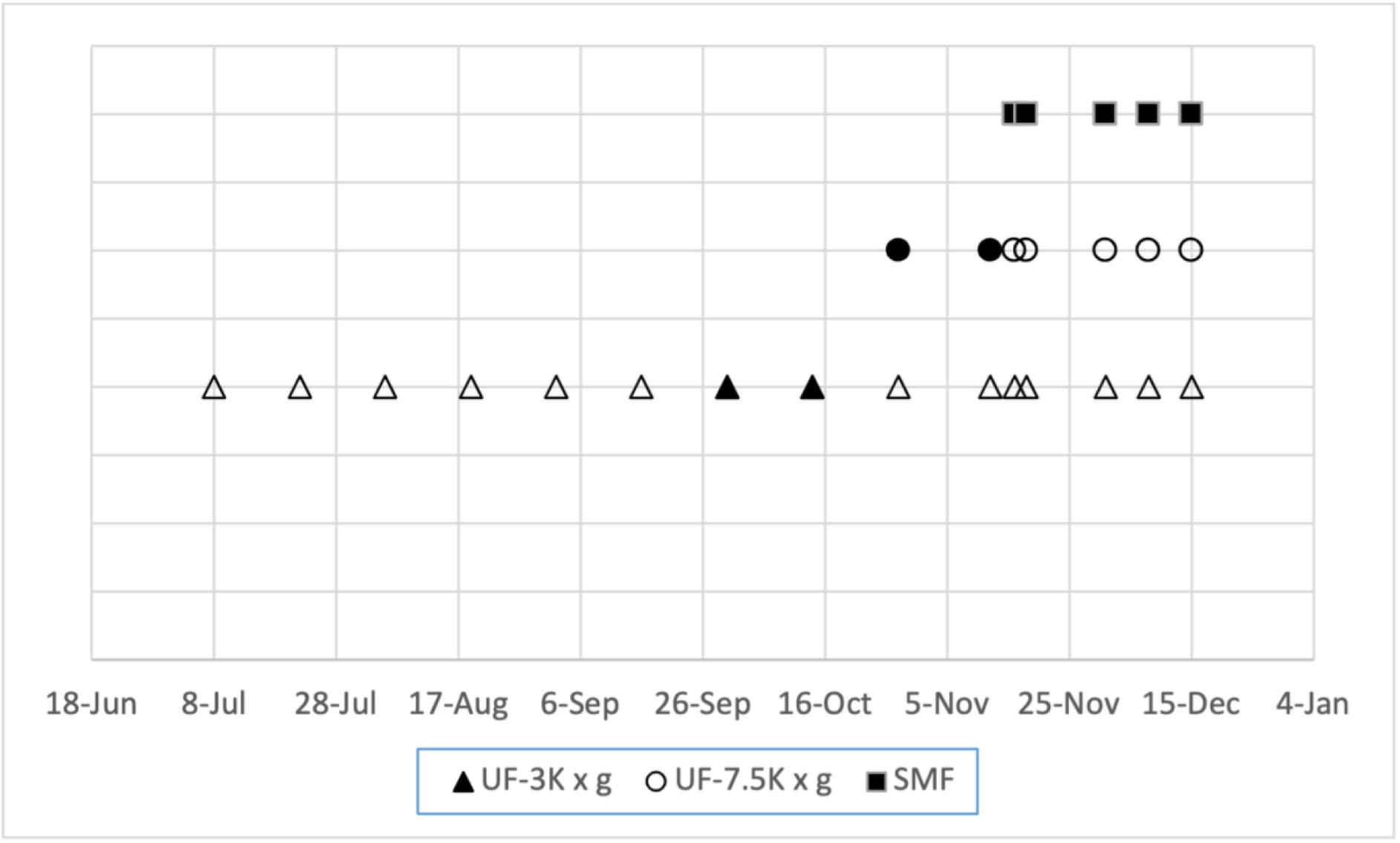
Detection of SARS-CoV-2 RNA in influent samples with different concentration methods. Filled shapes represent the samples that are SARS-CoV-2 positive, i.e. C_T_ is <40.

Non-detection of SARS-CoV-2 with UF-3K x g and UF-7.5K x g methods can be attributed to several factors, such as degradation during transportation and storage of samples (Ahmed et al., 2020b; Bivins et al., 2020a; Hokajärvi et al., 2021a; Hokajärvi et al., 2021b), losses during virus concentration (Chik et al., 2021; Ye et al., 2016), losses during extraction, and PCR related issues, such as inhibition and incomplete reverse transcription (Bustin et al., 2009). Since the variables in this study were the storage period at 4°C and virus concentration methods, the first two factors could explain the variations in detection with different concentration methods. SARS-CoV-2 was detected with UF-3K x g method only on September 30^th^ and October 14^th^ (Fig. 2). All samples processed 4 days after collection were negative, possibly due to decay of SARS-CoV-2 at 4°C (Ahmed et al., 2020b; Hokajärvi et al., 2021a) and partitioning of SARS-CoV-2 to the solid phase rather than liquid phase (Chik et al., 2021). The time required for the decay of 90% (T_90_) of N1 and N2 genes at 4°C was reported as 27.8 ± 4.45 and 36 days by Ahmed et al. (2020b) and Hokajärvi et al. (2021a). Considering that T_90_ values being much greater than four days, the decay of SARS-CoV-2 might not have lowered SARS-CoV-2 concentration in influent samples to undetectable levels. Moreover, Markt et al. (2021) reported a minimal reduction in SARS-CoV-2 concentrations in 9 days when stored at 4°C. Therefore, attachment of SARS-CoV-2 to solids, concentration method, and variation in recovery efficiency are plausible causes for the non-detection of SARS-CoV-2. Several studies have reported a higher affinity of enveloped viruses (mouse hepatitis virus [MHV] and *Pseudomonas* phage Φ6) to attach solid particles in wastewater compared to non-enveloped viruses (Ye et al., 2016), which might be the case for the non-detection of SARS-CoV-2 in influent filtrates in this study. Attachment of SARS-CoV-2 to solids and variations in recovery efficiencies together possibly resulted in non-detection. This was further confirmed by the analysis of solid particles retained on the filters. SARS-CoV-2 was detected in all solids retained on the filters except for the sample collected from WESTP on October 14^th^ (Table 4). The difference of one order of magnitude between SARS-CoV-2 concentrations in solids and filtrate indicated the significance of solid phase partitioning of SARS-CoV-2. For some samples, SARS-CoV-2 concentration was even higher in solids. Hokajärvi et al. (2021a) also reported a higher detection frequency of SARS-CoV-2 N2 gene frequency in solids (89%) compared to that in the supernatant (67%). In most cases, the virus might predominate in solid phase (Chik et al., 2021). Similarly, some other studies also emphasized solids-associated behavior of SARS-CoV-2 by taking the absorption of viral particles to solids into consideration and reporting the correlations between virus concentration and case data (D’Aoust et al., 2021; Graham et al., 2021; Peccia et al., 2020). Effects of freezing and thawing on SARS-CoV-2 concentrations in solids should also be considered in this study. We extracted from solids on filters stored at - 20°C that were later subjected to freeze-thaw once. Markt et al. (2021) reported a significant SARS-CoV-2 reduction in influent samples subjected to one freeze-thaw cycle. In comparison with these results, Simpson et al. (2021) and Hokajärvi et al. (2021a) observed a relatively lower reduction of SARS-CoV-2 in solids after one cycle of freeze-thaw, but still up to 60% of reduction. Considering current literature, the actual SARS-CoV-2 concentrations in solids might be higher than the measured concentrations. Moreover, the presence of solids in wastewater might enhance the stability of SARS-CoV-2 (Kitajima et al., 2020).

**Table 4.**
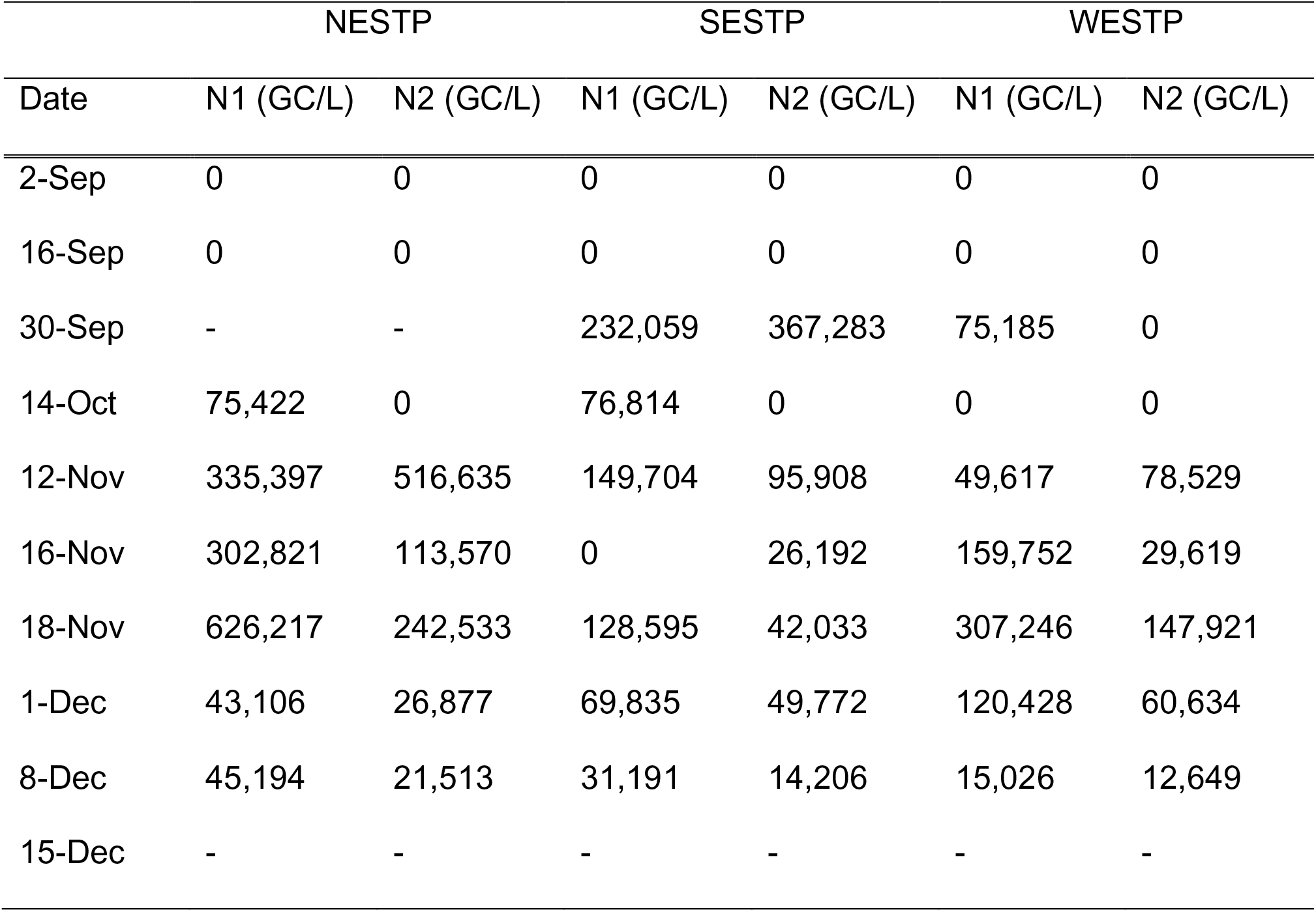
SARS-CoV-2 RNA concentration in influent solids from NESTP, SESTP and WESTP.

Samples processed with UF-7.5K x g exhibited similar behavior to samples processed with UF-3K x g. SARS-CoV-2 was detected with this method only on October 28^th^ and November 12^th^ (Fig. 2 and Table 3) in influent filtrates. As discussed for UF-3K x g, retention of solids during prefiltration with 0.45 and 0.2 μm filters might result in the non-detection of SARS-CoV-2 in influent samples. In addition to the elimination of solids particles, increasing the centrifugation speed to 7500 x g might have led to the passage of viruses through the filter and/or binding of viruses onto filters. Indeed, the inability of recovery tests for virus concentration methods in determining the true recovery efficiencies can cause biases in the detection and quantitation of viruses in wastewater samples. Since SARS-CoV-2 is predominantly in contact with fecal matter in the wastewater matrix during the transportation to the WWTPs via sewer systems, recovery efficiency determination approaches, which are generally based on a spike-and-recovery approach, might not sufficiently reflect the actual recovery values of SARS-CoV-2 (Chik et al., 2021). This could be due to several reasons in the current study. First, there were only 30 minutes between sample spiking and processing in this study. Secondly, physicochemical characteristics of the spiked wastewater samples could be significantly different from influent samples that were processed in this study. Amoah et al. (2022) previously demonstrated the effects of pH, ammonia, and total solids on viral detection and SARS-CoV-2 concentration. Thus, the actual recovery of SARS-CoV-2 might be much lower than the assessed recovery efficiencies for UF-3K x g and UF-7.5K x g considering spike-and-recovery approaches and variations in physicochemical characteristics of the samples. Furthermore, higher concentrations of SARS-CoV-2 were observed in solids (Table 4) when total solids concentrations were higher in the samples collected from NESTP between October 28^th^ and November 18^th^ (Table S4). This increase can be attributed to the increase in total solids due to the partitioning of SARS-CoV-2 to solid phase (Chik et al., 2021). The relationship between case data (Fig. 3) and SARS-CoV-2 concentration in solids during this period should not be overlooked, as discussed in section 3.2.4.

**Fig. 3.**
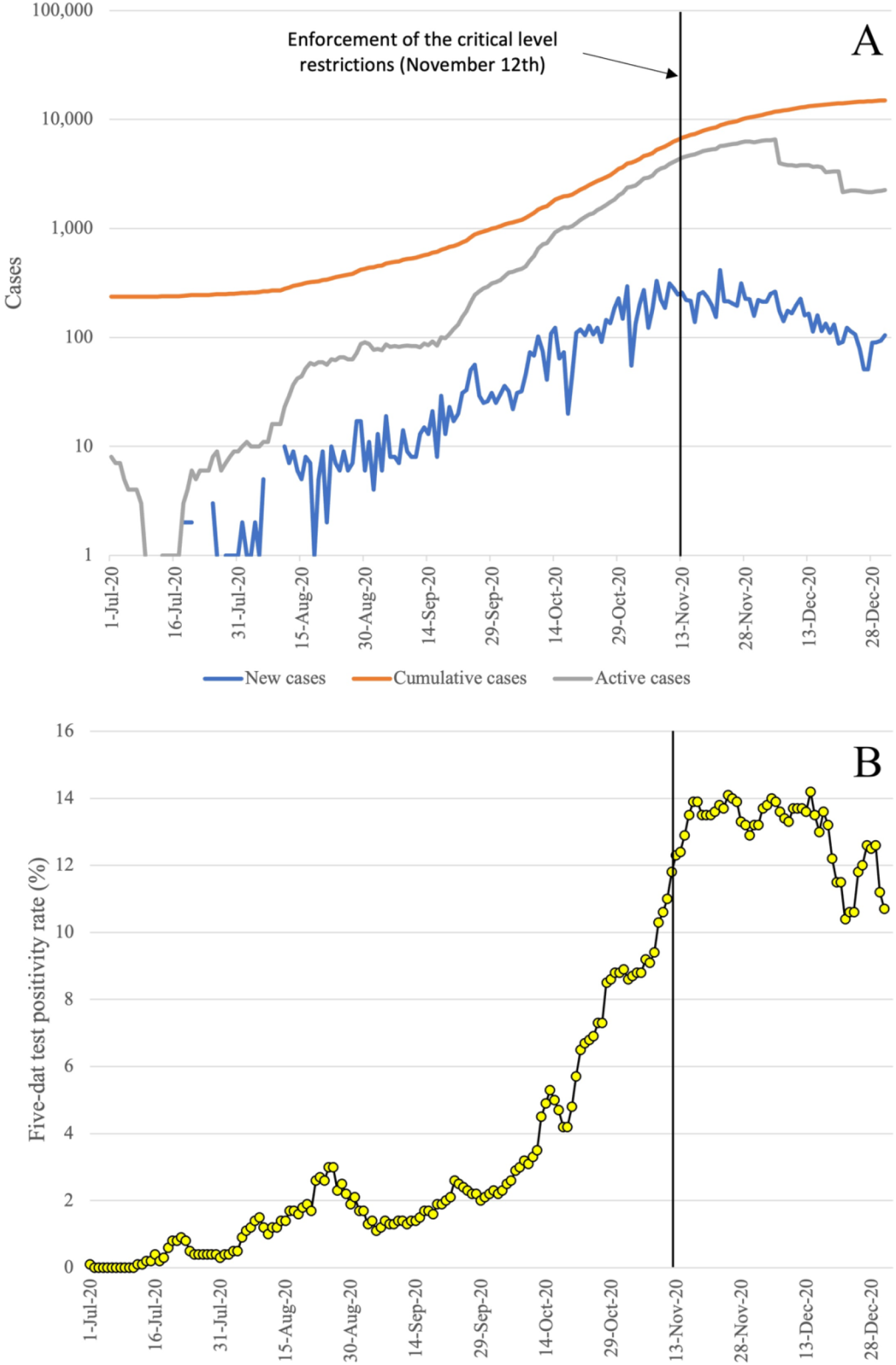
COVID-19 infection dynamics in Winnipeg. A) Temporal changes in the numbers of new, active, and cumulative cases during sampling period. B) Five-day test positivity rates during the sampling period.

SARS-CoV-2 was not detected between November 16^th^ and December 15^th^ when samples were processed with UF-3K x g and UF-7.5K x g methods. However, SARS-CoV-2 was detected in these samples using SMF (Fig. 2 and Table 3), although the sample processing was delayed between 46 and 75 days (Fig. 2). The successful detection of the virus was most probably because of the inclusion of both solids and supernatant into sample processing with SMF. The positive results confirmed that SARS-CoV-2 might predominate in solid phase and have higher stability in wastewater when attached to solids. An inter-laboratory study processing wastewater samples collected on the same day from a WWTP in Winnipeg also reported that SARS-CoV-2 was detected only by the laboratories processing samples with the methods focusing on solids (Chik et al., 2021). Another study on the partition of enveloped viruses to wastewater solids strengthens the claim of such solid-associated behavior with concentrations of enveloped viruses in solids found 1000 times higher than those in influent on a per mass basis (Ye et al., 2016). As a matter of fact, many other studies have suggested that a wastewater solids-based measurement of SARS-CoV-2 might be a more sensitive approach than a wastewater supernatant-based one (D’Aoust et al., 2021; Graham et al., 2021; Peccia et al., 2020; Westhaus et al., 2021). Moreover, input volumes of SMF (500 mL), UF-3K x g (120 mL) and UF-7.5K x g (120 mL) might be another factor determining detection of SARS-CoV-2, especially when the virus is not abundant in the samples.

SARS-CoV-2 can be detectable in both influent wastewater and solids long after storage at 4°C (Ahmed et al., 2020b; Hokajärvi et al., 2021a; Markt et al., 2021). The time required for 90 % reduction of SARS-CoV-2 in influent wastewater at 4°C was reported to be between 28, 36, and 52 days for the genomic targets, N1, N2 and E, respectively (Ahmed et al., 2020b; Hokajärvi et al., 2021a). On the other hand, only a one order of magnitude reduction of SARS-CoV-2 in solids was observed for the samples stored at 4°C over 100 days (Markt et al., 2021). SARS-CoV-2 was detected at higher frequencies and concentrations in solids compared to those in wastewater supernatant at the end of a period of 84 days (Hokajärvi et al., 2021a). Although the studies on the stability and detection of SARS-CoV-2 in influent and solids are limited, the current findings point out the predominant partition of SARS-CoV-2 to solid phase and higher stability of SARS-CoV-2 in solids. The results of this study, together with previous studies (Chik et al., 2021; Hokajärvi et al., 2021a; Markt et al., 2021), underscores the importance of the inclusion of both solids and supernatant fractions in wastewater processing for the detection of SARS-CoV-2. However, adsorption capacity of SARS-CoV-2 to wastewater solids needs to be investigated.

#### 3.2.2. Detection in primary sludge samples

Both N1 and N2 genomic targets of SARS-CoV-2 were detected in all primary sludge samples collected from NESTP and SESTP while they were not detected on November 16^th^, and only N2 was detected on November 18^th^ and December 1^st^ in the samples collected from WESTP (Table 3). The input volume of primary sludge samples processed for the detection of SARS-CoV-2 in this study was only 300 μL, which is much lower than the input volumes in previous studies (Graham et al., 2021; Peccia et al., 2020). Still SARS-CoV-2 was detected, indicating the high density of SARS-CoV-2 in primary sludge.

Detection of SARS-CoV-2 in primary sludge samples could be associated with the input volume and total solids in the samples. Low input volumes can cause biases in the detection and quantification of SARS-CoV-2 when virus concentration in the sample is low, and the samples are not homogenous in terms of total solids and SARS-CoV-2 distribution. Considering the high affinity of enveloped viruses to attach to the solids particles (Ye et al., 2016), total solids concentration can be another determinant of detection and quantification of SARS-CoV-2 in primary solids in addition to the community infection dynamics. In fact, concentration of SARS-CoV-2 was generally the highest in the samples collected from NESTP with an average primary sludge density of 3.53 ± 0.84 % and was generally the lowest in the samples collected from WESTP with an average sludge density of 0.37 ± 0.06 % (Table S5). This statement assumes homogeneous distribution of prevalence and incidence throughout the city of Winnipeg on the sampling days based on the high correlation between influent viral concentrations in WWTPs (r>0.91 for N1 and r>0.72 for N2) (Table S6). Higher input volumes should be considered for consistent and sensitive detection and quantification of the viral RNA in sludge samples.

#### 3.2.3. Detection in effluent samples

SARS-CoV-2 was not detected in both secondary and final effluent samples of all three WWTPs (Table 3), although it is worth mentioning that WESTP employs solar disinfection, which effectively reduces bacteria and is questionable in removing viruses, especially in cold climate conditions (Parsa et al., 2021). Employing UV disinfection, NESTP and SESTP might have effectively reduced SARS-CoV-2 concentration below detectable levels in final effluent. SARS-CoV-2, being an enveloped virus, is expected to be more sensitive to disinfection processes (UV, ozonation, and chlorination) than non-enveloped viruses (Chen et al., 2021; Saawarn and Hait, 2021). Non-detection of SARS-CoV-2 in tertiary-treated effluent samples was also reported elsewhere (Randazzo et al., 2020; Sherchan et al., 2020), indicating the efficacy of disinfection processes in the removal of SARS-CoV-2 considering high positivity rates (>83 %) in influent samples. This claim was further supported by the occurrence of SARS-CoV-2 in secondary-treated effluent (before disinfection) samples (Haramoto et al., 2020; Randazzo et al., 2020).

SARS-CoV-2 is removed not only by disinfection processes but also primary and secondary treatment processes, which can also effectively reduce SARS-CoV-2 concentration in the WWTPs (Balboa et al., 2020; Randazzo et al., 2020; Sherchan et al., 2020). Detection of SARS-CoV-2 in very low volumes of primary sludge samples in this study might indicate a significant removal of SARS-CoV-2 with primary treatment. While removal of SARS-CoV-2 with primary, secondary and tertiary treatments could explain non-detection of SARS-CoV-2 in effluent samples to a great extent, elimination of solid particles with prefiltration of effluent samples with 0.45 and 0.2 µm filters could be another factor considering the attachment of SARS-CoV-2 to solids. However, more studies are needed to fully uncover the fate of SARS-CoV-2 in WWTPs and the effects of virus concentration methods and input volume on the detection and quantification of SARS-CoV-2 in effluent samples.

#### 3.2.4. Detection of SARS-CoV-2 relates to the number of cases and test positivity rate

SARS-CoV-2 was first detected at a concentration of <50 gene copies per liter (GC/mL) on September 30^th^ in all influent wastewater samples collected from NESTP, SESTP, and WESTP (Table 3) when the number of the reported new cases and active cases in Winnipeg were 25 and 321, respectively (Fig. 3). This first detection concentration on September 30^th^ went up to 400 GC/mL after the concentration of the viral RNA in the solids was added to the concentration in supernatant. Detection of SARS-CoV-2 in wastewater samples collected from a WWTP in Winnipeg on August 31^st^ was first reported by Chik et al. (2021) at trace concentrations (<20 GC/mL), with the concentration methods focusing on solid fraction when number of the reported new and active cases were 11 and 85, respectively. Both results suggest a larger number of new and active cases consisting of asymptomatic, presymptomatic, and recovering cases of COVID-19 in the community. This is consistent with the estimations of the actual number of infections being 3 to 20 (Wu et al., 2020) and 6 to 24 times (Havers et al., 2020) higher than reported cases in the U.S. A study by Hong et al. (2021) focusing on the estimation of the minimum number of SARS-CoV-2 infected cases for the detection of viral RNA in wastewater estimated a minimum number of active cases of 253 to 459 positive cases per 10,000 population to detect SARS-CoV-2 in wastewater, which is much higher than the number of active cases on the day of the first detection in Winnipeg considering the population of Winnipeg (766,900) (Winnipeg, 2021). However, the reported range of active cases needed for the detection of SARS-CoV-2 might be a system- or site-specific estimation since the detection of SARS-CoV-2 in wastewater depends on many factors, including sampling frequencies, concentration methods, and their recovery efficiencies, RT-qPCR detection performance, sample size, daily wastewater flow rates (dilution factor), and environmental factors which can affect the persistence and abundance of SARS-CoV-2 in wastewater (Hong et al., 2021). Moreover, new cases predominantly contribute to the concentration of SARS-CoV-2 in wastewater rather than active cases (Gerrity et al., 2021; Wu et al., 2022) due to higher viral shedding rates in the very early stages of the infection (Benefield et al., 2020; He et al., 2020; Wei et al., 2020; Wölfel et al., 2020). Therefore, the number of active cases, without the data regarding new and early-stage cases, may not be enough in determining the minimum number of cases to detect SARS-CoV-2 in wastewater.

The difference between symptom onset and test confirmation suggests that time-lag might be another explanation for the detection of SARS-CoV-2 when the number of active cases was low. There is also a typical 4 to 5 day incubation period before onset of the symptoms (He et al., 2020; Li et al., 2020). A possible effect of time-lag can be further confirmed by the sharp increase in the number of cases and five-day test positivity rate (Fig. 3) after September 30^th^. Between the first sampling date, July 8^th^, and September 30^th^, the five-day test positivity rate fluctuated between 0.0% and 3.0 % (Fig. 3) (Manitoba, 2021). On September 30^th^, the test positivity rate was 2.1 %, and henceforth a constant increase in the test positivity rate until mid-December was noticed, with a peak test positivity rate of 14.2 % on December 14^th^. During this period, we detected SARS-CoV-2 in all influent samples (including solids fractions) collected from three wastewater treatment plants, except on October 28^th^ (Table 3), using different concentration methods.

Previous studies have reported associations between the concentration of SARS-CoV-2 in primary settled solids and COVID-19 cases (Graham et al., 2021; Peccia et al., 2020). In this study, SARS-CoV-2 was detected in almost all primary sludge samples. In the previous sections, the fluctuations in the concentration of SARS-CoV-2 were partly associated with total solids in the samples. A direct correlation between concentrations and reported cases is intentionally avoided due to low-resolution sample collection and small input volumes of primary sludge. These two factors cause some difficulties in justifying the fluctuations in SARS-CoV-2 concentrations in primary sludge samples. In other words, it is not clear if solids, the number of infections, or a combination of them cause fluctuations in SARS-CoV-2 concentrations. High-resolution sample collection (at least weekly) and larger input volumes could generate more reliable and consistent results for a wastewater surveillance study.

### 3.3. Estimation of Concentrations and Shedding Cases

Using reported decay rates for N1 and N2 gene fragments (Ahmed et al., 2020b; Hokajärvi et al., 2021a), initial concentrations of SARS-CoV-2 in the late processed influent samples, collected between November 16^th^ and December 15^th^, were estimated and then converted into normalized concentration using eqn. 1. Reported COVID-19 infection dynamics and observed (measured and back-trajectory modeled) concentrations were compared with infection dynamics and generated by model 2 as an attempt to justify models and understand the course of COVID-19 infection dynamics in the city. Lack of information regarding population and COVID-19 infection dynamics in each sewershed is one of the limitations of this study, preventing us from doing a sewershed-based analysis.

Peak concentrations of SARS-CoV-2 were observed on the day when critical-level restrictions were enforced and the following 4^th^ and 6^th^ day (Fig. 4). Other observed concentrations after the first detection were generally in close proximity with the modeled concentrations for at least one genomic target. Different decay rates for N1 and N2 resulted in higher concentrations of N1 for back-trajectory modeled samples collected between November 16^th^ and December 15^th,^ while the difference between N1 and N2 concentrations for the rest of the samples was smaller than 0.35 on log_10_ scale (Fig. 4). The decay rate of N1 was calculated based on the degradation of genomic signals of gamma-irradiated SARS-CoV-2 by Ahmed et al. (2020b) and had a higher standard deviation of 15% and lower R^2^ of 0.79 compared to those values of N2, which were based on degradation of active SARS-CoV-2 (Hokajärvi et al., 2021a) (Table 2). Gamma irradiation of SARS-CoV-2 and relatively higher variations in the decay rate of N1 might result in significant biases in such a back-trajectory modeling approach. Therefore, we mostly consider N2 concentrations for back-trajectory modeled samples in the discussion.

**Fig. 4.**
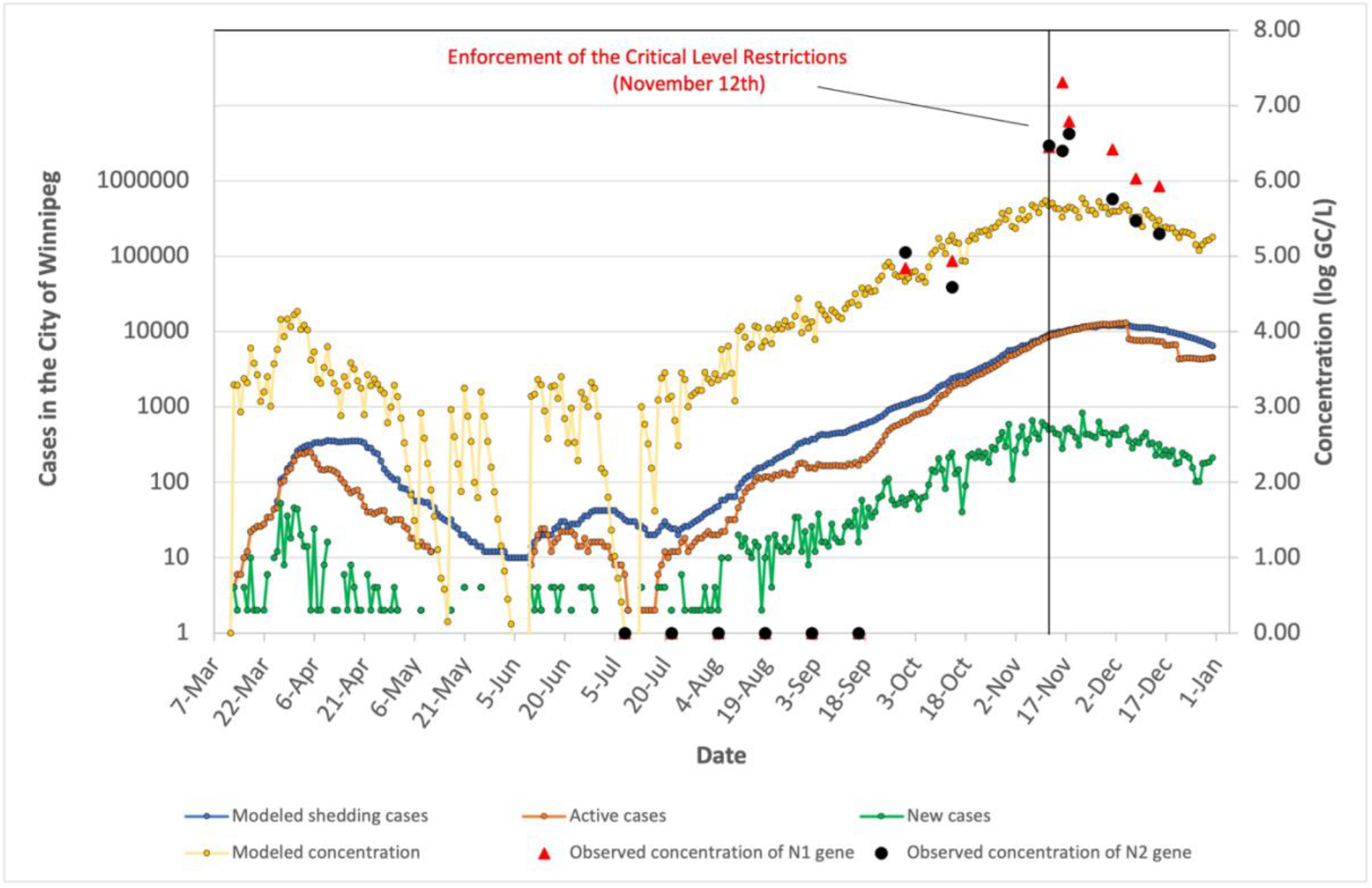
Modeled (yellow circles) and observed concentrations (red triangles and black circles) of SARS-CoV-2 RNA as log10 concentrations and COVID-19 infection dynamics from the beginning of the outbreak in Winnipeg. Observed concentrations between September 30^th^ and November 12^th^ are the sum of the concentration in solids and filtrates since the latter concentrations were obtained using SMF which concentrates viruses from both supernatant and solids. Observed concentrations between November 16^th^ and December 15^th^ are estimated using eqn. 3. Observed concentrations are represented for both N1 (red triangles) and N2 (black circles). Green circles represent new cases in Winnipeg corrected with an ascertainment ratio of 2. Oranges circles represent active cases in Winnipeg corrected with an ascertainment ratio of 2. Blue circles represent modeled shedding cases in Winnipeg, i.e. modeled active cases.

The critical level restrictions in Manitoba were applied when extensive community transmission of COVID-19 occurred, outbreaks could not be controlled, and a heavy toll on the health-care system is expected. With the enforcement of the restrictions on November 12^th^, gatherings and travels were extremely restricted, and schools and non-essential businesses were closed. Bearing in mind that the outbreak and transmission are at the critical levels when the restrictions are applied, the occurrence of a much higher number of new cases compared to reported numbers is most likely because of the limited testing capacity at the peak times and time-lag between the incubation period of SARS-CoV-2, onset of the symptoms and test confirmation (He et al., 2020; Li et al., 2020). In this study, such a scenario was validated with the observed concentrations of SARS-CoV-2 in influent that peaked following the enforcement in November and significantly lowered to the modeled concentrations in December as the outbreak was contained. The difference between the observed concentrations of N2 and modeled concentrations (0.79 to 0.97 on log_10_ basis) in November suggests that the actual number of cases might be much higher than reported numbers, which is further supported by the sharp increase in test-positivity rates and the number of new cases (Fig. 3). These findings correspond to the findings of Havers et al. (2020), reporting that the actual number of cases can be as high as 24 times of reported numbers.

In general, observed concentrations were sensitive to the fluctuations in test-positivity rates and the number of new cases except for the samples collected in November. Model 2 assumed an ascertainment ratio of 2, which is an optimistic assumption considering ascertainment ratios up to 24 in the literature (Havers et al., 2020; Wu et al., 2020). While an ascertainment ratio of 2 in the model generated comparable concentrations with the observed concentrations for September, October, and December samples, a higher ascertainment ratio might come into question during the peak times in November. The high concentrations and ascertainment ratios in November can be explained by the high transmission rates of the disease and the number of asymptomatic and presymptomatic cases around this period. After the enforcement of critical level restrictions, public mobility and disease transmission were expected to be minimized. However, the number of new cases and the test-positivity rate remained high during the sampling in November and December, most likely due to time-lag associated with the peaking of viral load and disease transmissivity prior to symptom onset (Benefield et al., 2020; He et al., 2020; Wei et al., 2020). Probably most individuals were infected around the first day of the enforcement but developed symptoms later, which was in agreement with the typical incubation period of 4-5 days before symptom onset (Guan et al., 2020; Lauer et al., 2020; Li et al., 2020), and only sought healthcare and underwent testing after symptom onset. SARS-CoV-2 load in December might be due to prolonged shedding from individuals infected earlier. Additionally, individuals infected before the enforcement can infect other people living with them in subsequent days, and disease transmission can still be high among essential workers and their families after the enforcement.

Estimation of SARS-CoV-2 concentration based on shedding rate and reported cases has been previously studied with an emphasis on the significant contribution of newly infected individuals to SARS-CoV-2 loads in wastewater and occurrence of the highest fecal shedding rates in the first days of infection before symptoms develops and tests are conducted (Gerrity et al., 2021; Wu et al., 2022). They also reported high similarity between the observed and modeled concentrations, which validates the efficacity of such models to estimate concentration and infection dynamics. Sensitivity of SARS-CoV-2 levels in wastewater to the fluctuations in the number of new cases and test-positivity rates in this study suggest that an initial burst of viral shedding may occur in the very early stages of COVID-19 infection even before symptom onset and may be followed by a prolonged period of lower shedding rates up to 30 days as reported by Chen et al. (2020), Hoffmann and Alsing (2021), Wölfel et al. (2020) and (Wu et al., 2022).

Our data suggest that a back-trajectory model as a function of decay rate and time may be used to estimate initial concentrations of late processed samples as it fit the modeled concentrations and fluctuations in new cases. High-resolution sampling and site-specific decay rates may help to validate and improve the model. Time-lag, testing capacity, and test-positivity rates in addition to the reported cases and shedding rate should also be considered for the shedding rate-based models to obtain better model fits and estimations.

## 4. Conclusions

Wastewater surveillance of COVID-19 has been considered an early-warning tool for potential outbreaks and an informative method to characterize COVID-19 infection dynamics. However, researchers have not reached a consensus on sample collection and processing, and data analysis. Furthermore, the potential discharge of SARS-CoV-2 from wastewater treatment plants to the environment is another issue that requires a comprehensive investigation of the fate of SARS-CoV-2 throughout the wastewater treatment process.

The current study focused on the detection of SARS-CoV-2 in wastewater and sludge samples and the relationship between detection and infection dynamics in Winnipeg. Our results showed that SARS-CoV-2 might predominate in solids. Concentration methods focusing on supernatant and solids fractions may perform better in virus recovery, especially for enveloped viruses. Thus, the type of concentration method may significantly affect SARS-CoV-2 recovery from influent samples.

SARS-CoV-2 might be substantially removed during primary and secondary treatment as SARS-CoV-2 was detected in influent and primary sludge but not in secondary and final effluents. The high affinity of SARS-CoV-2 to solids and detection of SARS-CoV-2 in primary sludge samples at high concentrations suggest sludge line as a potential removal mechanism and a sampling spot for wastewater surveillance. Improvement in processing sludge samples for viral concentration and detection is required to gain more insight into the fate of SARS-CoV-2 in sludge line and optimize sampling and processing.

In addition to the detection and fate of SARS-CoV-2 in WWTPs, the proposed study underlines the relationship between SARS-CoV-2 levels in influent samples and infection dynamics characterized by increasing COVID-19 incidence and prevalence during the sampling period. Both observed and modeled concentrations were sensitive to the fluctuations in new cases and test-positivity rates, suggesting an early burst of viral shedding in infected individuals. During the peak times, the number of infections can be much higher than the number of reported cases considering the time-lag between infection and test confirmation, and asymptomatic infections. To confirm our findings and improve such shedding rate-based models, additional studies with higher sampling resolution and the models informed by some other factors factors, such as time-lag, test capacity, and test-positivity rates, are required. Overall, this study demonstrates that SARS-CoV-2 may predominate in solids, and wastewater surveillance of COVID-19 can provide valuable insights into infection dynamics prior to clinical test confirmations.

## Data Availability

All data produced in the present work are contained in the manuscript

## Acknowledgments

We thank the City of Winnipeg, the management, and operators at the NESTP, SESTP, and WESTP for collecting samples and providing information regarding wastewater characteristics, in particular Jong Hwang, Brendan Hellrung, Mark Lefko, Jorge Martins, and Shaun Walker. We acknowledge Ms. Shana Mann and Miss Jocelyn Zambrano (University of Manitoba) for nucleic acid extraction from solids. We also thank Tri Le (University of Manitoba) for proofreading.

## CRediT authorship contribution statement

**Kadir Yanaç:** Conceptualization, Methodology, Validation, Formal Analysis, Investigation, Data Curation, Visualization, Writing & Original Draft,. **Adeola Adegoke:** Software, Formal Analysis. **Liqun Wang:** Software, Formal Analysis. **Qiuyan Yuan:** Conceptualization, Methodology, Validation, Resources, Review & Editing, Supervision, Project Administration, Funding Acquisition. **Miguel Uyaguari:** Conceptualization, Methodology, Validation, Resources, Review & Editing, Supervision, Project Administration, Funding Acquisition.

