## Supplementary material for "Detection of SARS-CoV-2 RNA Throughout Wastewater Treatment Plants and A Modeling Approach to Understand COVID-19 Infection Dynamics in Winnipeg, Canada": Supplementary.docx

**Title Page**

**Author’s names:** Kadir Yanaç ^1^, Adeola Adegoke^2^, Ligun Wang^2^, Qiuyan Yuan^1^ Miguel Uyaguari^3^

^1^ Department of Civil Engineering, University of Manitoba, Winnipeg, Manitoba, Canada

^2^ Department of Statistics, University of Manitoba, Winnipeg, Manitoba, Canada

^3^ Department of Microbiology, University of Manitoba, Winnipeg, Manitoba, Canada

**Table S1.** Sampling dates and sample types collected from three wastewater treatment plants.

| Sampling date | 24-h Composite Wastewater Samples | Primary Sludge Samples | Secondary (Before) and Final Effluent Samples (After disinfection) |
| --- | --- | --- | --- |
| July 8, 2020 | X |  | X |
| July 22, 2020 | X |  | X |
| August 5, 2020 | X |  | X |
| August 19, 2020 | X |  | X |
| September 2, 2020 | X |  | X |
| September 16, 2020 | X |  | X |
| September 30, 2020 | X |  | X |
| October 14, 2020 | X |  | X |
| October 28, 2020 | X |  | X |
| November 12, 2020 | X | X |  |
| November 16, 2020 | X | X |  |
| November 18, 2020 | X | X |  |
| December 1, 2020 | X | X |  |
| December 8, 2020 | X | X |  |
| December 15, 2020 | X | X |  |

**Table S2.** Key properties of WWTPs sampled

| WWTP | Treatment Process | Average Daily Flow Treated (million liters per day) | Population Served |
| --- | --- | --- | --- |
| NESTP | HPO followed by UV disinfection | 195 | 404,000 |
| SESTP | HPO followed by UV disinfection | 58 | 176,000 |
| WESTP | A2O followed by natural light disinfection in the summer and cooling in the winter | 20 | 86,000 |

HPO: High purity oxygen wastewater treatment, A2O: Sequential anaerobic-anoxic-oxic wastewater treatment

**Table S3.** Primer/probe sets of RT-qPCR assays

| **Assay** | **Primer/Probe** | **Concentration** | **Sequence** |
| --- | --- | --- | --- |
| N1 | 2019-nCoV_N1-F | 200 nM | 5’-GACCCCAAAATCAGCGAAAT-3’ |
|  | 2019-nCoV_N1-R | 200 nM | 5’-TCTGGTTACTGCCAGTTGAATCTG-3’ |
|  | 2019-nCoV_N1-P | 200 nM | 5’-FAM-ACCCCGCATTACGTTTGGTGGACC-ZEN/Iowa Black-3’ |
| N2 | 2019-nCoV_N2-F | 200 nM | 5’-TTACAAACATTGGCCGCAAA-3’ |
|  | 2019-nCoV_N2-R | 200 nM | 5’-GCGCGACATTCCGAAGAA-3’ |
|  | 2019-nCoV_N2-P | 200 nM | 5’-FAM-ACAATTTGCCCCCAGCGCTTCAG- ZEN/Iowa Black-3’ |
| Armored RNA | Arm_RNA-F | 400 Nm | 5’- AGCCTGTCAATACCTGCACC-3’ |
|  | Arm_RNA-R | 400 Nm | 5’- CACGCTTAGATCTCCGTGCT-3’ |
|  | Arm_RNA-P | 200 Nm | 5’ Cy5-AGAGTATGAGAGGTCGACGA-TAO 3’ |

**Table S4.** TS and TSS in influent samples of NESTP, SESTP, WESTP

|  | **NESTP** | | **SESTP** | | **WESTP** | |
| --- | --- | --- | --- | --- | --- | --- |
| **Date** | **TS (mg/L)** | **TSS (mg/L)** | **TS (mg/L)** | **TSS (mg/L)** | **TS (mg/L)** | **TSS (mg/L)** |
| 8-Jul | 974 | 220 | 1,272 | 202 | 958 | 204 |
| 22-Jul | 1,160 | 264 | 924 | 300 | 984 | 192 |
| 5-Aug | 1,070 | 320 | 802 | 202 | 1,100 | 208 |
| 19-Aug | 1,050 | 315 | 798 | 200 | 884 | 206 |
| 2-Sep | 644 | 280 | 614 | 114 | 842 | 231 |
| 16-Sep | 1,000 | 304 | 756 | 192 | 982 | 208 |
| 30-Sep | 1,030 | 320 | 812 | 268 | 868 | 236 |
| 14-Oct | 1,070 | 372 | 822 | 240 | 804 | 200 |
| 28-Oct | 1,620 | 455 | 872 | 233 | 830 | 243 |
| 12-Nov | 1,520 | 1,080 | 806 | 231 | 768 | 207 |
| 16-Nov | 1,490 | 1,450 | 718 | 200 | 726 | 177 |
| 18-Nov | 1,600 | 843 | 754 | 197 | 808 | 323 |
| 1-Dec | 1,110 | 350 | 760 | 200 | 772 | 217 |
| 8-Dec | 1,100 | 307 | 810 | 233 | 844 | 253 |
| 15-Dec | 1,000 | 210 | 860 | 280 | 814 | 236 |

**Table S5.** Primary Sludge Densities in NESTP, SESTP, WESTP

|  | Primary sludge densities Density (%) | | |
| --- | --- | --- | --- |
|  | NESTP | SESTP | WESTP |
| 12-Nov | 2.42 | 3.42 | 0.38 |
| 16-Nov | 4.32 | 2.24 | 0.31 |
| 18-Nov | 2.83 | 1.94 | 0.29 |
| 01-Dec | 4.1 | 2.29 | 0.34 |
| 08-Dec | 3.11 | 2.68 | 0.42 |
| 15-Dec | 4.37 | 2.44 | 0.45 |

**Table S6.** Correlation between SARS-CoV-2 RNA concentrations in NEST, SESTP, WESTP. N1) Correlation for N1. N2) Correlation for N2

| *N1* | *NESTP* | *SESTP* | *WESTP* |
| --- | --- | --- | --- |
| NESTP | 1 |  |  |
| SESTP | 0.989748233 | 1 |  |
| WESTP | 0.913370328 | 0.928222603 | 1 |
| *N2* | *NESTP* | *SESTP* | *WESTP* |
| NESTP | 1 |  |  |
| SESTP | 0.72753165 | 1 |  |
| WESTP | 0.921271604 | 0.875798662 | 1 |

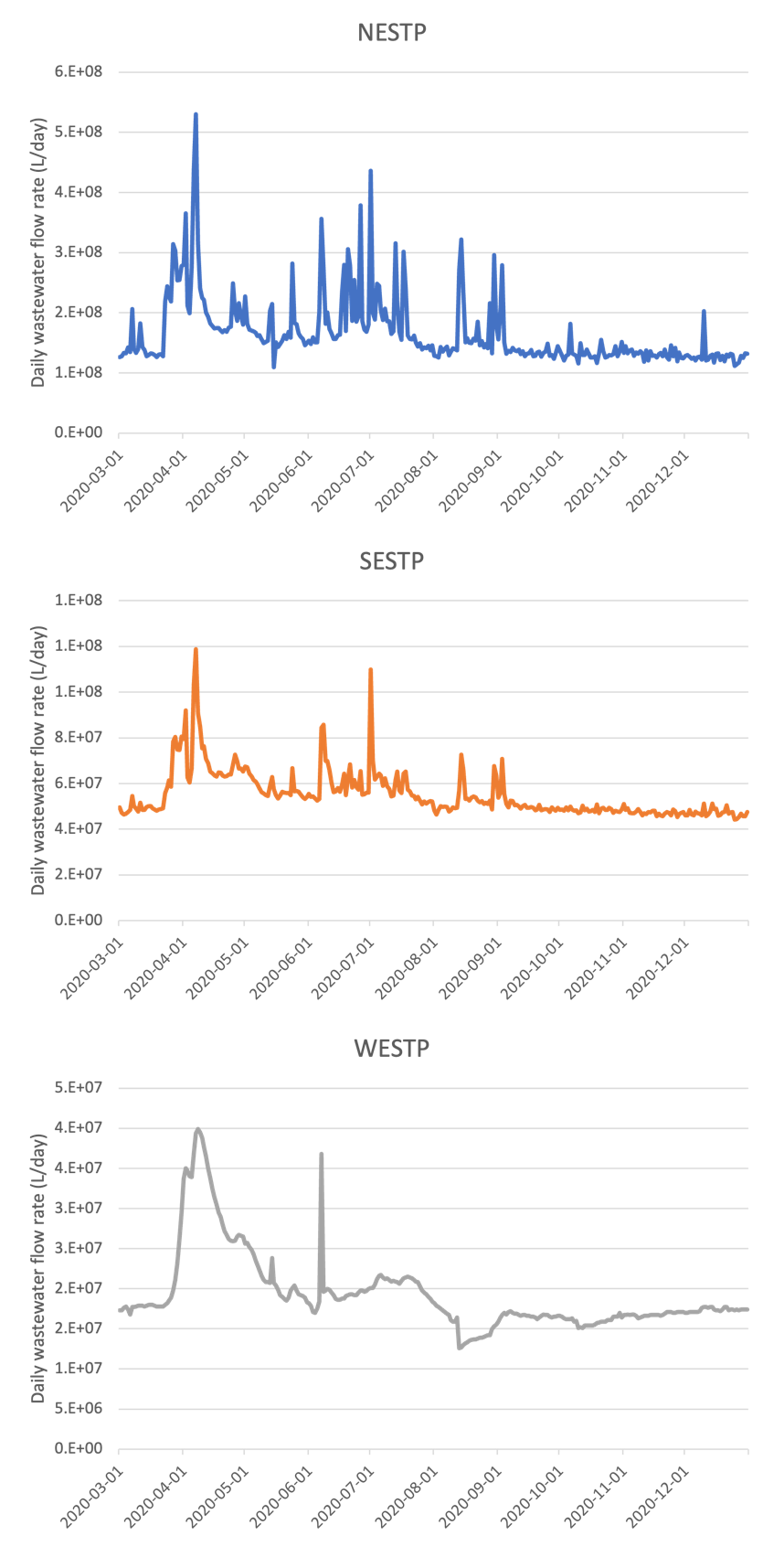

**Fig. S1.** Daily wastewater flowrates in NESTP, SESTP and WESTP

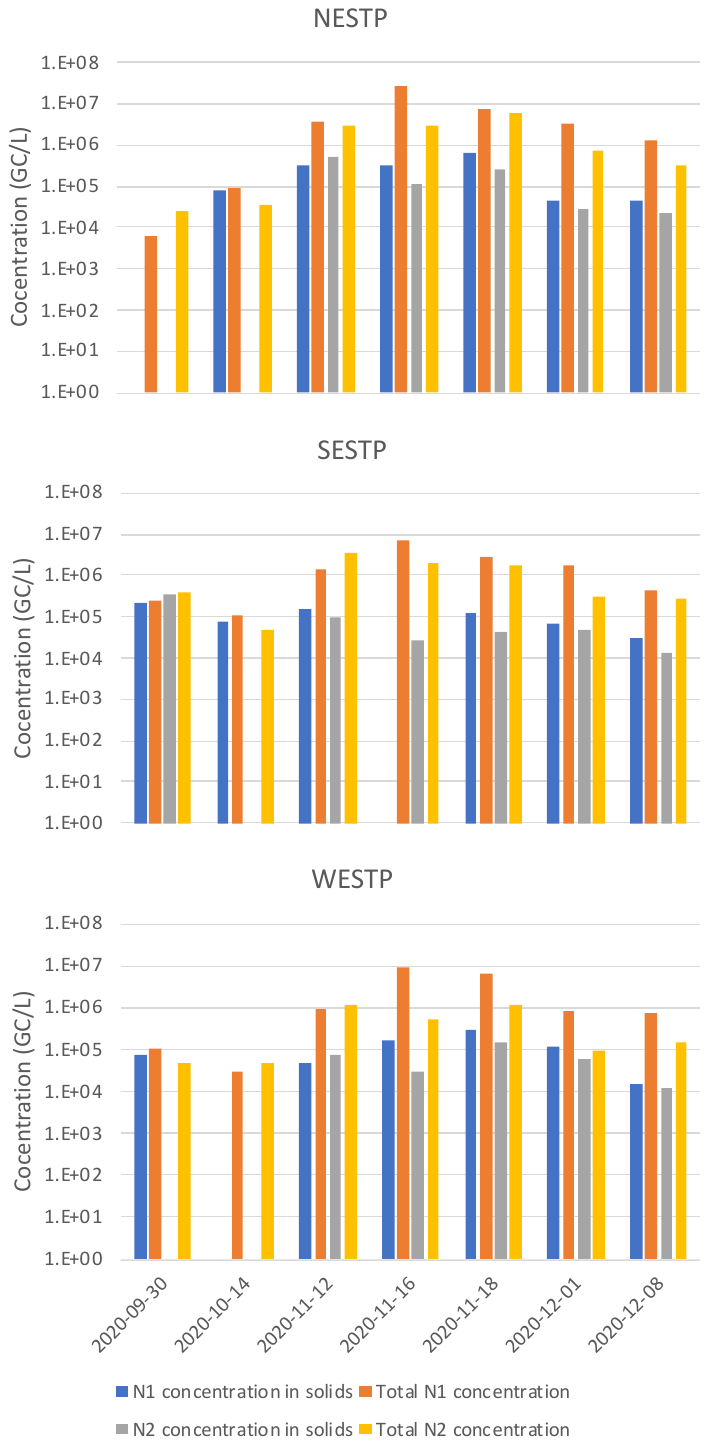

**Fig. S2.** Concentrations of N1 and N2 in solids and inluent samples. Total concentration is the sum of virus concentration in solids and filtrates.
